# High COVID-19 incidence among Norwegian travellers returned from Lombardy: implications for travel restrictions

**DOI:** 10.1101/2020.03.20.20038406

**Authors:** Ola Brynildsrud, Vegard Eldholm

## Abstract

On February 27th, three cases of COVID-19 were reported among Norwegians that had recently returned from Lombardy, Italy. Travellers from the region rapidly became the most common source of imported infections in the earliest stage of the Norwegian COVID-19 epidemic. The situation was exacerbated by the unfortunate temporal overlap between the Norwegian winter holidays and intense epidemic spread of COVID-19 in Northern Italy, resulting in a large number of infected travellers. Here we combined flight data on travels between Norway and Lombardy with patient-level data to determine the fraction of travellers returning to Norway that had been infected with SARS-CoV-2.

Travellers returning to Norway from Lombardy contracted COVID-19 at incidence rates up to 0.02 per person-day in the period spanning February 21st and March 1st, with a clear uptick in transmission in the middle of the period.

This shows an example of the infection risk in tourist destinations being several fold higher than elsewhere in the region. In Norway, this is also supported by high rates of infections among tourists returning from Austria in February and March, despite a low number of reported cases in the country at the time.

The massive COVID-19 prevalence among travellers suggest that mandatory quarantine of returning travellers or suspension of non-essential international flights is essential if the aim is to control or suppress the COVID-19 pandemic.

## Results and Discussion

In Fig. 1, daily numbers of diagnosed COVID-19 cases who had recently returned from Lombardy are shown together with the temporal distribution of passengers flying in each direction. We find that ∼3% of travellers who returned to Norway from Lombardy between February 21st and March 1st were eventually diagnosed with SARS-CoV-2 infection. We find a clear increase in the fraction of infected travellers during the period, from ∼1% on Feb 21st-25th with an exponential increase to ∼9% on March 1st (Fig. 2). Similarly, we calculate the incidence rate per person-day among Norwegians in Lombardy to be roughly 3.4e-3 on Feb 21st-25th, up to 0.02 per person-day on March 1st.

**Figure 1.**
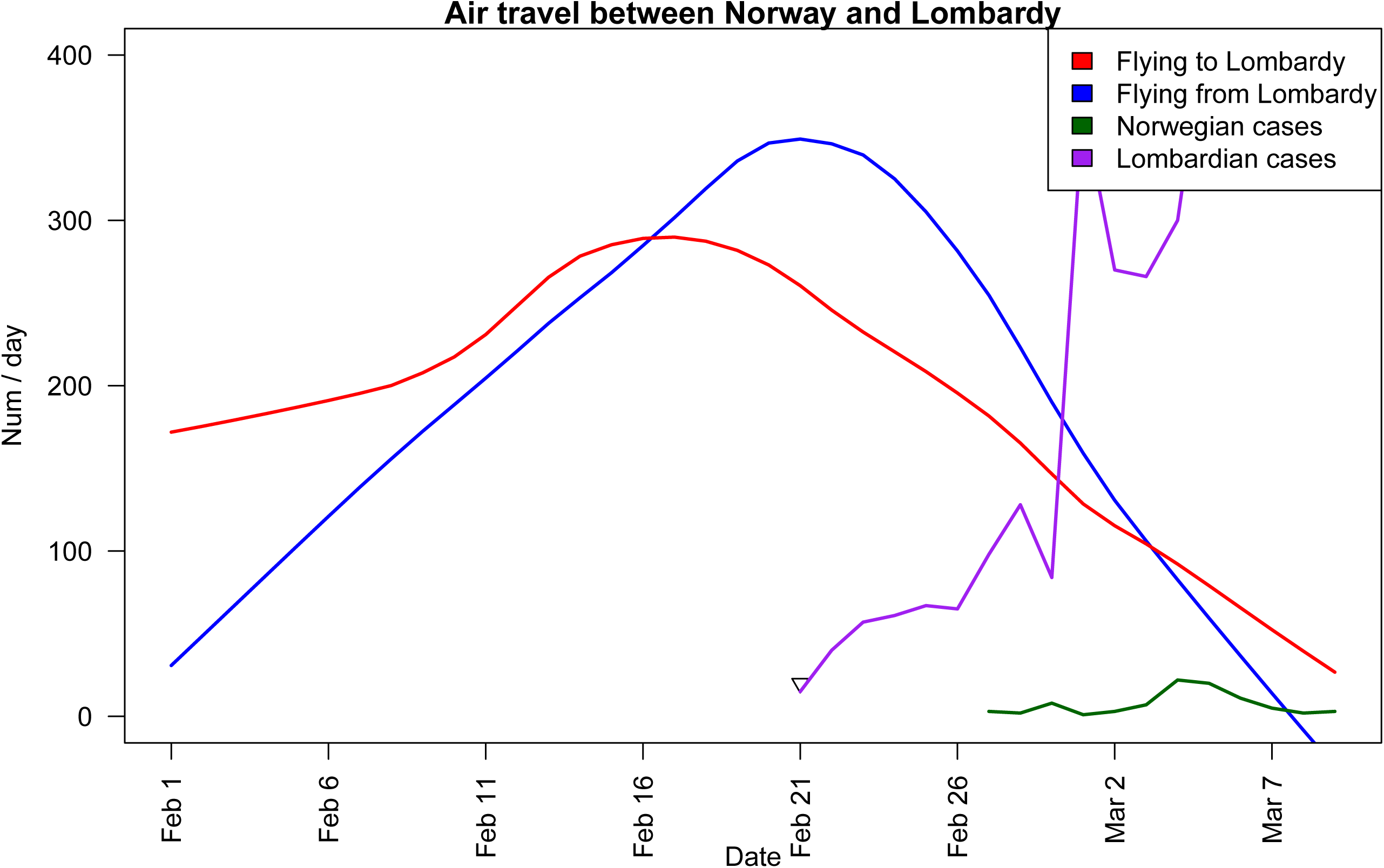
Air-travel between Norway and Lombardy, with reported cases in Norway among travellers returning from Lombardy. The green bars indicate confirmed cases in Norway linked to travel in Italy. The first 14 cases in Lombardy were detected on Feb 21st. The Norwegian winter vacation lasts for one week, but the exact week varies between regions. In total the vacation was spread over three weeks spanning February 21st - March 6th.

**Figure 2.**
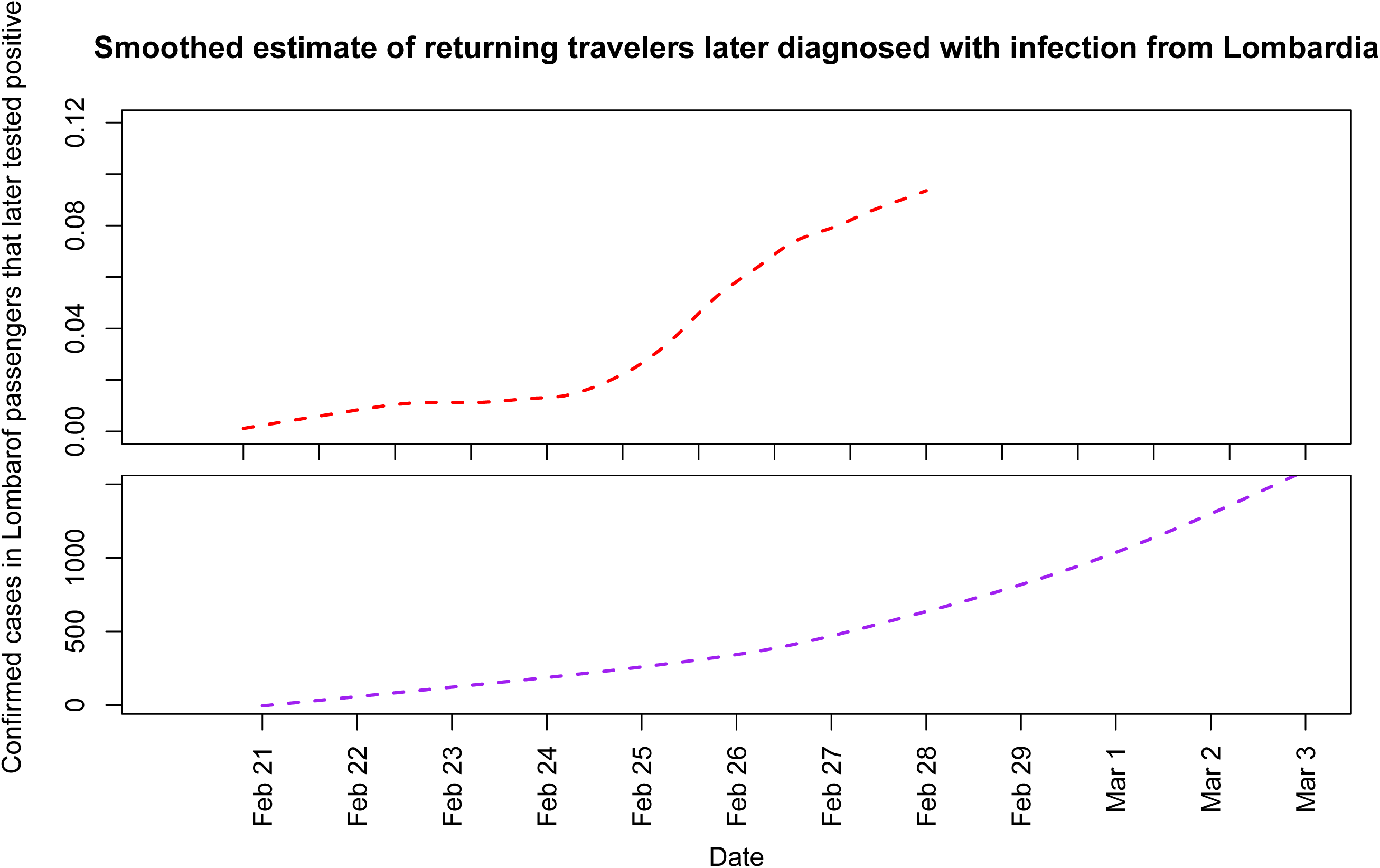
COVID-19 infection-load among travellers returning from Lombardy. (Top) Estimated fraction of flight passengers returning to Norway infected with Sars-CoV-2. (Bottom) Cumulative number of notified cases in the Lombardy region.

These results confirm that COVID-19 infections were contracted at very high rates by travellers to Lombardy in late February.

This period overlapped with the Norwegian winter vacation, and it is known that the vast majority of travellers were tourists heading for the Italian alps and ski resorts. In the same time-period, the number of daily reported cases in the Lombardy region rose from 15 to 369 in a population of ∼10,000,000.

As the number of diagnosed cases in Lombardy in the early days of the epidemic is clearly not representative of the true COVID-19 prevalence at the time, we used daily numbers of fatalities to estimate the true case numbers at the time. Based on the number of deaths reported in Lombardy on March 13th (146), and assuming an average of 20 days (Jung et al. 2020) from infection to death for fatal cases and a case fatality ratio of 1% (Famulare 2020; Liu et al. 2020), we estimate that the true number of new cases in Lombardy on February 22nd was ∼14600, compared to 40 reported cases. This rough estimate suggests that as little as 1% of cases were detected.

The pronounced uptick in infections diagnosed among passengers returning after February 25th is difficult to reconcile with the growth of the local epidemic (Fig. 2). This might indicate that the infection-pressure increased quite abruptly at one or more tourist hubs such as ski-resorts or other localities. The very high infection rates seen among travelers returning to Norway indicates that tourist hubs might have been important early epicenters of the infection in Europe.

## Conclusion

Travellers returning to Norway from Lombardy contracted COVID-19 at extreme rates in the period spanning February 21st and March 1st, with a clear uptick in transmission in the middle of the period.

The infection risk associated with international air travel is impossible to predict, as reported cases in a region might be only a fraction of the true case load, and also because the infection pressure in tourist destinations might be several fold higher than the overall pressure in a region. In Norway, this is also supported by extreme rates of infections among tourists returning from Austria in February and March, despite a low number of reported cases in the country at the time.

## Methods

### Data collection

Case statistics from the Lombardy region of Italy was collected from the official git of the Civil Protection department of Italy - Presidenza del Consiglio dei Ministeri - Dipartimanto della Protezione Civile (https://github.com/pcm-dpc/COVID-19).

Flight data was kindly contributed by Avinor - a state-owned limited company that operates most civil airports in Norway. The data included all commercial direct flights between any Lombardian airport (all of which were Milanese) and an Avinor airport in the period Feb 1st - Mar 10th. The data also included passengers leaving an Avinor airport towards Milano with an indirect (“connecting”) flight, but not those arriving from Lombardy via other airports. The latter was inferred by assuming equal proportions of direct vs indirect travel on flights to and from Lombardy. Notably, the data didn’t include information about non-Avinor-operated airports. This means that flights from Torp/Sandefjord airport are not included. A maximum of two flights per week were flying between Torp and Milano in the study period, and among patients that were diagnosed in Norway, none were reported to have been on this route. Travel by alternative means such as by car or train were assumed to be too uncommon to matter in our estimates.

Patient-level information about Norwegian SARS-CoV-2 cases as well as epidemiological tracking was contributed by the Norwegian Institute of Public Health.

### Estimates using returning travelers

Daily passenger-level travel counts were smoothed with a lowess function using a smoother span of 0.4. Daily number of confirmed cases in Norway was smoothed with a span of 0.66. From contact tracing of the patients that had been to Lombardy we established that they had returned by flight ∼5 days prior to receiving a positive diagnosis. However, since this information was not available for all cases, we proceeded as if all cases had returned home exactly 5 days before diagnosis. Based on contact information as well as previous estimates of incubation period (Lauer et al. 2020; Li et al. 2020) and time from symptom onset to diagnosis (Jung et al. 2020), we assumed that they had been infected in Lombardy. For person-time, we assumed that all travelers stayed in Lombardy for four days.

### Ethical considerations

All data used in the current study was anonymized prior to being obtained by the authors. Flight statistics was aggregated and could not be used to identify individuals. Daily number of cases by municipality with epidemiological origin is in the public domain and posted daily in reports from the Norwegian Institute of Public Health.

## Data Availability

Data refered to in the manuscript is available at https://github.com/admiralenola/lombardy-norway

## Notes

### Competing Interest Statement

The authors have declared no competing interest.

### Funding Statement

No external funding was received

